# Integrating respiratory infection surveillance and temperature data improves all-season mortality reconstruction

**DOI:** 10.64898/2026.08.15.26359980

**Authors:** Junyu Wang, Matthias an der Heiden, Christopher Irrgang

## Abstract

All-season mortality surveillance can inform public-health planning under climate change, but attribution is complicated by overlapping effects of temperature, respiratory infections and other non-environmental factors. Here we develop a component-based neural-network model for daily all-cause mortality that combines high-resolution temperature and dew-point data, severe acute respiratory infection (SARI) hospitalization incidence, demographic structure and an adaptive mortality baseline. In Germany, inputs included non-COVID SARI and COVID-19-associated SARI. Using 2014–2025 mortality data, the hybrid model reduced daily root-mean-square error to 80.2 deaths, compared with 170.2 for a weather-only model and 117.1 for an infection-only model, and enabled age- and sex-resolved examination of fitted components. Including respiratory-infection indicators reduced the increase in risk assigned by the model to cold exposure, suggesting that temperature-only models may partly assign winter infection-related variation to cold. Component magnitudes represent conditional model decompositions rather than causal estimates of deaths due to SARI or temperature.

## Introduction

Understanding and modeling mortality is fundamental for public-health planning, risk assessment and policy implementation. Accurate mortality mod-eling supports mortality projections and informs decisions on health-system capacity, demographic change, pandemic preparedness and emergency response.

It is important not only to track aggregate mortality but also to attribute mortality variation to specific drivers over time. Although death certificates are well documented in many countries, cause assignment often depends on situational judgment [1, 2]. Certificates can list multiple causes, but burden-of-disease analyses typically use only the underlying cause, while contributory or triggering conditions are rarely counted [3, 4]. As a result, the impact and interplay of exposures such as respiratory infections, heat, and cold cannot always be measured directly from registered causes of death. Statistical attribution methods address this limitation by estimating mortality associated with specific exposures, which is relevant for climate change, demographic change, and pandemic preparedness.

Statistical frameworks such as generalized additive models (GAMs) [5] and distributed lag non-linear models (DLNMs) [6] are widely used to quantify temperature-associated mortality. These models capture delayed and non-linear associations between temperature and mortality, often identifying a minimum-mortality temperature range and attributing non-optimal temperature exposures to excess deaths [7]. However, two limitations remain important for temperature-only models. First, humidity modifies heat stress and may influence cold-associated risk [8, 9, 10], yet there is no universally accepted index for summarizing combined temperature–humidity effects, and many studies omit or simplify humidity [11, 12]. Second, cold-associated mortality may be overestimated when winter mortality peaks are attributed mainly to temperature. For example, Europe experienced similar winter temperature anomalies in January 2016 (–1.2 *^◦^*C) and January 2017 (–1.3 *^◦^*C) [13], but mortality differed by 23.4% (470,992 vs. 581,329 deaths across 28 EU countries) [14]. Such differences point to additional winter mortality drivers, particularly respiratory infections.

Baseline estimation is itself part of this attribution problem. Conventional mortality baselines often either retain part of the seasonal mortality structure or rely on supposedly low-exposure reference periods that are not actually exposure-free [15]. This is particularly problematic in all-season analyses, where both winter and summer can contain substantial exposure-related mortality.

A key challenge for such all-season attribution is that respiratory infections exhibit strong seasonality and contribute to winter mortality in temperate regions [16], but their role is not fully represented in temperature-associated mortality models. Influenza surveillance systems exist in many European countries, but fewer countries maintain nationwide inpatient surveillance for severe acute respiratory infections (SARI), which are more closely linked to mortality than outpatient influenza-like illness reports [17]. After the COVID-19 pandemic, awareness of respiratory-infection surveillance increased, and several countries, including Germany, the Netherlands, and Portugal, expanded SARI monitoring [17, 18, 19]. However, available datasets remain limited in temporal depth and spatial resolution [20].

Together, these challenges leave few methodological approaches that examine all-season mortality and its environmental and infectious drivers in a unified framework. We address this gap with a component-based neural-network model that integrates SARI hospitalization data, COVID-19-associated SARI hospitalization data, high-resolution temperature and dew-point data, demographic structure, and an adaptive mortality baseline. Germany is used as a demonstration setting because the required data streams are available from 2014 onward. The primary objective is to improve reconstruction of all-cause and all-season mortality while separating temperature-associated, respiratory-infection-associated, and baseline components within the model framework. We therefore evaluate the framework mainly by reconstruction performance of observed all-cause mortality. These comparisons assess whether integrating environmental and respiratory-infection surveillance can improve all-season mortality monitoring, while the fitted components remain conditional model decompositions rather than causal estimates.

## Methods

### Data sources

The analysis dataset covered the period from October 13, 2014 to December 28, 2025.

### Mortality data

To define the outcome for model fitting and evaluation, we obtained all-cause mortality data from the Federal Statistical Office of Germany (Destatis) [21]. Three reporting formats were used:

1. Daily deaths by federal state and sex,
2. Weekly deaths by federal state, sex, and broad age groups (0–65, 65–75, 75–85, 85+),
3. Weekly deaths at national level by sex and detailed age groups (0–30, five-year age groups from 30–35 to 90–95, and 95+).

### Population data

Population data were used to calculate mortality rates and aggregate model outputs by demographic group. Population counts for Germany’s 400 administrative districts were obtained from the Destatis regional database up to and including 2024, stratified by five-year age group, from 0–5 to 90–95 and 95+, and by sex [22]. Annual counts were linearly interpolated to generate daily estimates for each district–age–sex subgroup. For subsequent dates, population values were held constant at their 2024 levels.

### Weather data

To represent environmental exposure, daily mean temperature and dew-point temperature from October 13, 2014 to December 28, 2025 were derived from the HOSTRADA high-resolution gridded climate dataset, provided by the German Weather Service (Deutscher Wetterdienst, DWD) [23]. HOSTRADA offers 1 *×* 1 km spatial resolution and hourly data for the study period. District-level time series were calculated as population-weighted averages, based on the 2022 census data [24] for the grid cells overlapping each district.

### Hospitalization data

To represent respiratory-infection pressure, national weekly hospitalization incidence for severe acute respiratory infections (SARI) and COVID-19associated SARI from October 13, 2014 to December 28, 2025 were obtained from the national SARI surveillance system operated by the Robert Koch Institute (RKI) [25]. A SARI hospitalization is defined as a hospital admission with an ICD-10 diagnosis code J09–J22 as the primary diagnosis. COVID-19-associated SARI is defined as a SARI case with an additional confirmed COVID-19 ICD-10 code U07.1. Non-COVID SARI incidence was obtained by subtracting COVID-19-associated SARI cases from total SARI incidence; it is hereafter referred to as “non-COVID SARI”. The dataset provides age- specific hospitalization incidence time series for the age groups 0–4, 5–14, 15–34, 35–59, 60–79, and 80+ years.

Influenza-specific SARI hospitalization incidence was additionally available from 2021 onward. This series is shown in Figure 1 for epidemiological context but was not included as a separate model input. Influenza-associated SARI therefore remained part of the aggregated non-COVID SARI incidence. Because SARI hospitalization data are available only at the national level, regional heterogeneity in infection pressure cannot yet be modeled. In the absence of regional SARI data, we assigned each district the same weekly age-specific national SARI incidence. Within each age band, incidence was assumed constant across constituent 5-year age classes (e.g., the reported 60–79 incidence was assigned to 60–64, 65–69, 70–74, and 75–79). Weekly values were converted to daily incidence series using monotone piecewise cubic Hermite interpolation.

**Figure 1.**
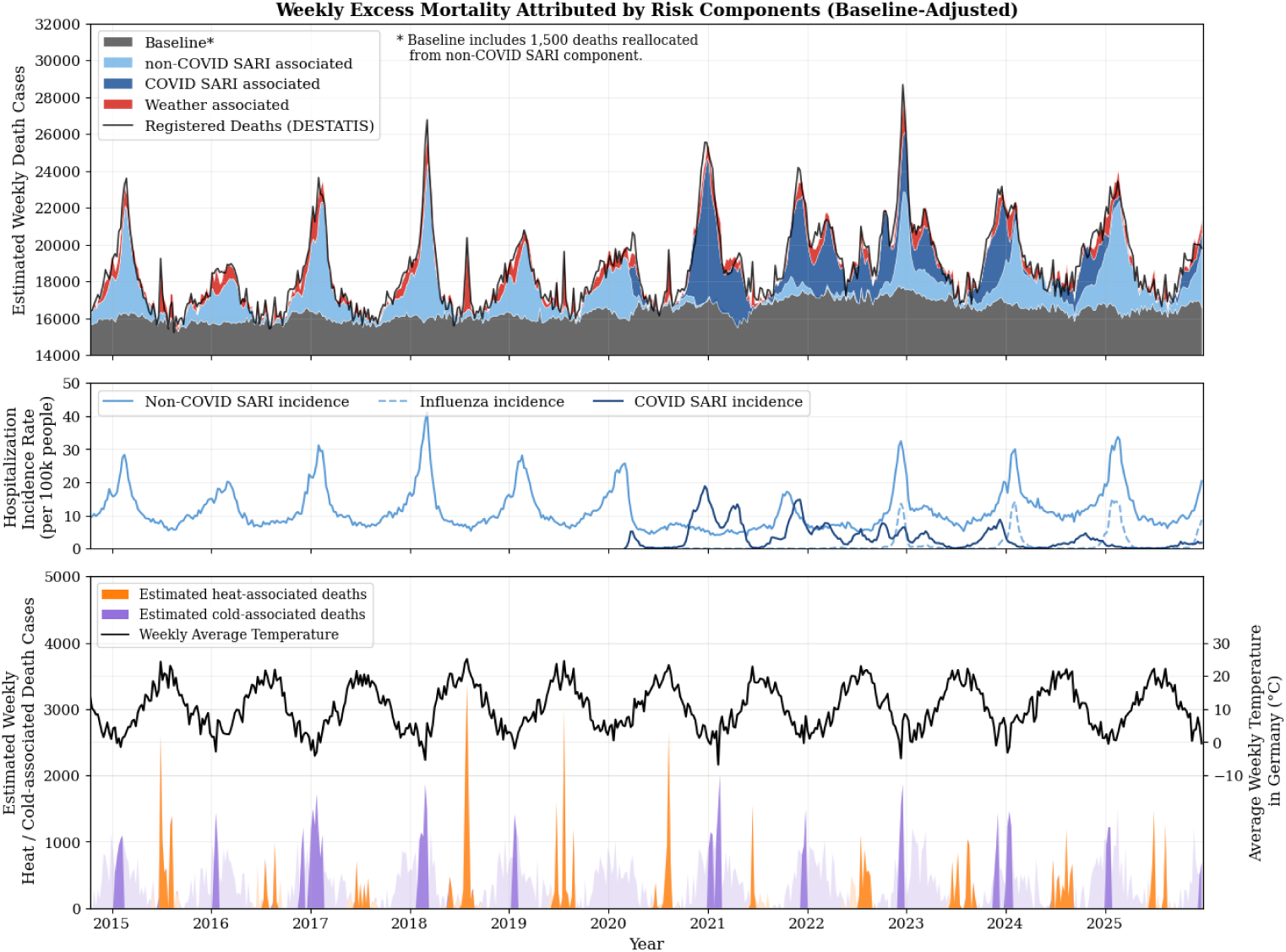
Component-based reconstruction of weekly mortality in Germany, 2014–2025. (A) Weekly mortality reconstructed from the fitted model components and baseline. Component-specific vertical offsets are used only for visual separation. (B) Weekly national hospitalization incidences for non-COVID SARI, COVID-19-associated SARI, and influenza. Influenza-specific SARI incidence is shown for contextual comparison only and was not modeled as a separate component. (C) Model-estimated temperature-associated deaths, separated into heat- and cold-associated components, shown with Germany-wide weekly average temperature. Highlighted weeks indicate average temperatures above 20 *^◦^*C or below 0 *^◦^*C in the corresponding week or adjacent weeks.

### Mortality Model Model overview

The described mortality, demographic, weather, and hospitalization data were combined in a component-based mortality model. The model extends our earlier approach for estimating heat-associated mortality [26] by adding respiratory-infection signals and an adaptive mortality baseline. Specifically, we introduce component-specific convolutional blocks for non-COVID SARI (abbreviated as nC-SARI in equations) and COVID-19-associated SARI, while retaining a baseline component that is not optimized by gradient descent but is periodically updated during training. We refer to the full configuration combining temperature, dew-point temperature, non-COVID SARI, COVID-19-associated SARI, and the adaptive baseline as the hybrid model. This joint temperature-, humidity-, and SARI-based feature design allows the model to reconstruct total mortality and to inspect potential winter misattribution in temperature-only models. A graphical illustration of the data flow and the training process is provided in Supplementary Figures 1 and 2. The model estimates daily all-cause death counts *m* for day *t*, district *d*, age group *a*, and sex *s*. Let *P_t,d,a,s_* denote the corresponding population and *b_t,d,a,s_* the corrected baseline mortality rate. Predicted deaths are obtained by multiplying the population by the baseline rate and the fitted relative-risk components:

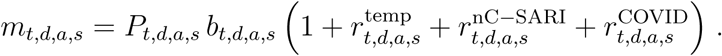

**Figure 2.**
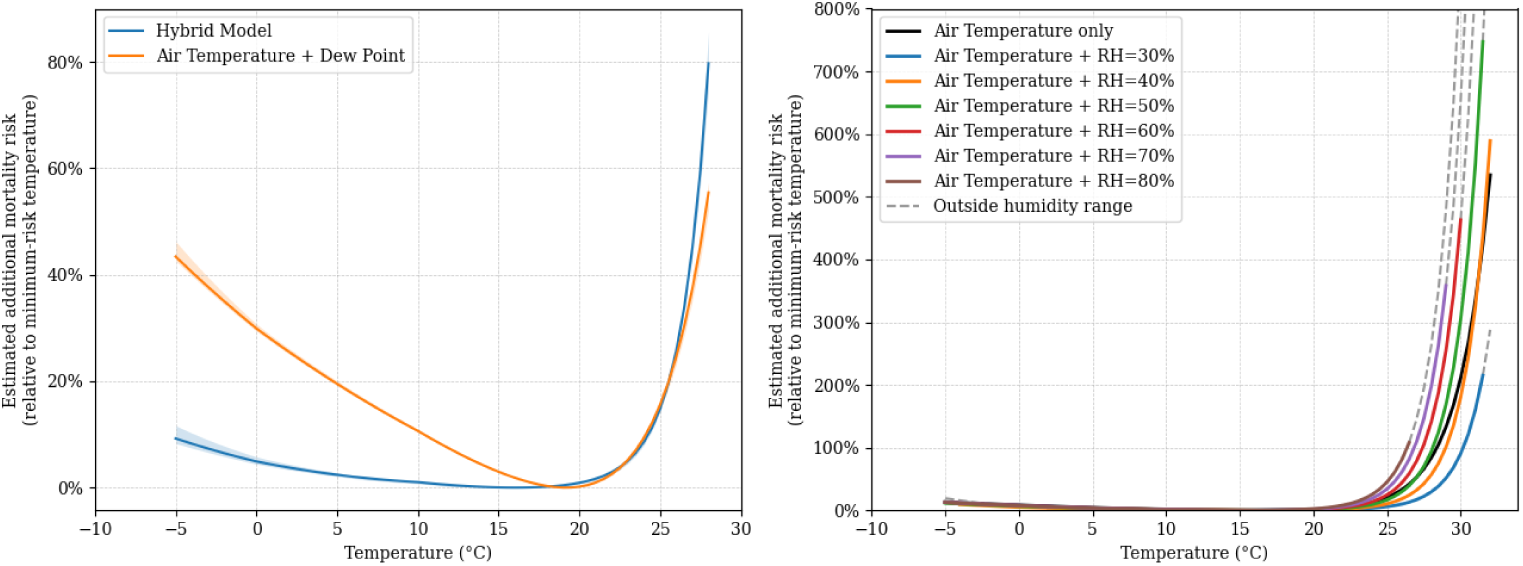
Modeled additional temperature-associated mortality risk under alternative model specifications and humidity conditions. All curves were calculated by setting the same air temperature and humidity conditions across the full 6-day weather input window. Left: additional mortality risk relative to the minimum-risk temperature for the hybrid model compared with a weather-only model using air temperature and dew-point temperature. Solid lines show the ensemble mean curves, and shaded areas indicate the minimum-to-maximum range across 10 ensemble members. Right: additional mortality risk for a model using air temperature only and for models using air temperature plus the dew-point humidity indicator under fixed relative humidity levels from 30% to 80%. Values are shown as (RR - 1) × 100, with 0 indicating no increase relative to the minimum-risk temperature. Dashed grey segments indicate temperature–humidity combinations outside the observed humidity range.

Component-attributable death estimates follow as:

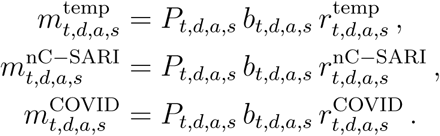

For readability, the equation is written in additive relative-risk notation. In the implementation, the weather block estimates a positive temperature-dependent mortality factor applied to the baseline, while the SARI and COVID-19 blocks estimate additional baseline-scaled mortality components. Model outputs are aggregated to the Destatis reporting resolutions before loss computation during the training procedure (see Model training and validation section below).

### Baseline mortality component

Baseline mortality represents the slowly varying background component of death records after removing the modeled short-term temperature- and respiratory- infection-associated variation. It is calculated separately for each demographic group (age and sex) and serves as a reference point for estimating excess mortality. It should therefore be interpreted as a model-based decomposition component rather than as a directly observed exposure-free mortality level.

In this study, the baseline is dynamically updated during training to account for changes in mortality patterns over time, including demographic shifts and other slow changes in background mortality. By incorporating updated residual information, the model provides a time-varying estimate of background mortality.

The dynamic baseline is updated every 50 epochs during model training (see sections below) using the following process: for each demographic group (age *a*, sex *s*), weekly residual deaths for Germany are computed as:

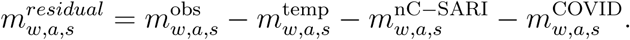

Here, 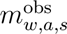 are the observed death counts from Destatis, and 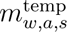, 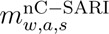, 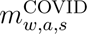 represent the model-predicted contributions from temperature, non-COVID SARI, and COVID-19-associated SARI, respectively.

For baseline adaptation, the temperature component used in the residual calculation is referenced internally to a neutral 16 *^◦^*C scenario. This internal reference is separate from the reported heat- and cold-associated attribution, which uses the category-specific counterfactual definitions described in the attribution section.

The residuals are then processed as follows:

- A median filter is applied to remove noise,
- An exponentially weighted moving average (EWMA) is used to smooth the data, and
- Linear interpolation generates daily estimates for the baseline, *b̂_t,a,s_*.

The primary single-sided smoother used only current and preceding residual observations and did not incorporate information from subsequent observations; the double-sided variant was evaluated only in sensitivity analysis. This method assumes that each district initially shares the same baseline mortality rate, which is then adjusted with separate correction factors. Notably, baseline parameters are not trainable and do not receive gradients during optimization.

### Temperature-associated mortality block

The temperature block estimates temperature-associated mortality relative to the baseline. It maps district-level daily mean temperature and dew point temperature (*T_t,d_, D_t,d_*) to 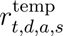 and accounts for lagged effects. Inputs pass through 1-D convolutional layers (kernel size 6, stride 1) over the current day and previous five days, selected by grid search, followed by an exponential activation and a fully connected layer to approximate the nonlinear temperature–mortality relationship commonly modeled with GAM and DLNM approaches.

### Non-COVID SARI-associated mortality block

The non-COVID SARI block estimates respiratory-infection-associated mortality relative to the baseline. Interpolated non-COVID SARI hospitalization incidence 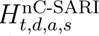 drives 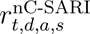 via a single 1-D convolution (kernel size 10, no bias) with non-negative weights so the attribution from incidence to mortality is always non-negative. When COVID-19-associated SARI is not modeled separately, the input combines non-COVID SARI and COVID-19- associated SARI incidence, *H*^nC-SARI^ + *H*^COVID^. Kernels are shared across the 16 federal states. The block is time-invariant, i.e., it does not impose seasonality or long-time trends on the non-COVID SARI effect.

### COVID-19-associated SARI mortality block

COVID-19-associated SARI incidence 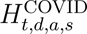 is processed in a separate pathway with the same structure as the non-COVID SARI block, yielding 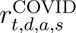. This separation allows distinct attribution for COVID-19-associated SARI and non-COVID SARI.

### Calibration and corrections

In addition to the exposure-related components, the model accounts for reporting and regional structure in the registered death records. Registered deaths exhibit systematic day-of-week variation (registration and certification workflows) and persistent state-level differences (socioeconomic factors). To account for these variations, we learn two multiplicative calibration terms on the baseline mortality: a correction factor *c*_1_ for the day of the week and a state offset *c*_2_. The corrected baseline is then

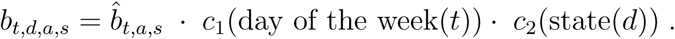

The temperature- and infection-attributable components are then calculated relative to this corrected baseline.

The state offset was handled separately because COVID-19 exhibited strong regional variation that is not captured in our inputs. Including pandemic-era mortality counts to train *c*_2_ would therefore distort state offsets. To avoid this, *c*_2_ is updated only using pre-COVID data (up to 2019), while *c*_1_ is trained on the full training period.

### Model training and validation

After defining the component blocks and calibration terms, the predicted death estimates *m_t,d,a,s_*were aggregated to match the registered death counts reported by Destatis, and model parameters were optimized using a Poisson deviance loss (See Supplementary Figures 2).

In preliminary experiments, optimization with the Poisson deviance alone tended to drift in the aggregate mean level and to favor mild overestimation of total deaths. To counteract this, we included a mean-alignment step during training. At each training step, an age- and sex-specific scaling factor was computed from the ratio of observed to predicted Germany-wide weekly deaths during the pre-COVID part of the training period, and this factor was used to define a mean-aligned training objective. During optimization, the mean-aligned target became increasingly important across epochs. All model comparisons reported in the main text and supplement used this mean-alignment procedure. Reported reconstruction performance and model-attributed component estimates were obtained from the trained 10-replicate model ensemble.

The model was trained using the AdamW optimizer. A ReduceLROnPlateau learning-rate schedule was applied based on training loss. Training was performed using full-batch updates without mini-batching to preserve temporal coherence in the daily time series. Model fitting was conducted on a single GPU.

Observations from October 13, 2014 to June 5, 2022 were used for training (75%), June 6, 2022 to December 29, 2024 served as the chronological validation period (25%), and data from 2025 were reserved as an additional test period. In period-specific analyses, the post-pandemic evaluation period refers to 2023–2025, combining the 2023–2024 validation years with the 2025 test period. Reconstruction performance on observed all-cause mortality is the primary validation target of the study. The component-specific outputs are then analyzed as interpretable latent contributions within the fitted model rather than as directly observed cause-specific death labels.

### Attribution of mortality components

After evaluating reconstruction performance, the fitted outputs can be decomposed to obtain model-attributed death estimates for each combination of day, district, age group, and sex. Death estimates attributed by the fitted model to temperature (*m*^temp^), non-COVID SARI (*m*^nC^*^−^*^SARI^), and COVID-19-associated SARI (*m*^COVID^) were derived from the fitted component terms and counterfactual calculations. Aggregation across spatial or demographic groups was performed by summing these model-attributed deaths across the relevant dimensions.

Model-attributed temperature-associated deaths were classified according to the daily temperature of each district, using an optimal-temperature range of 15–20 °C. Temperatures between 0 and 15 °C were classified as mild cold, while temperatures below 0 °C or above 20 °C were defined as cold and heat periods.

To estimate the mortality attributed by the fitted model to these categories, a counterfactual temperature curve was generated for each district. In this model, observed daily temperatures falling below the specific category’s lower bound were replaced with that bound, and those exceeding the upper bound were replaced with the upper bound (maintaining relative humidity). The resulting difference between the observed and generated temperature curves serves as the basis for calculating the excess mortality component for each temperature classification.

## Results

### Seasonal attribution of mortality components

Figure 1 shows weekly mortality reconstructed from the fitted model components from 2014 to 2025. The component-based reconstruction captured seasonal mortality patterns, including increases in winter mortality associated with respiratory-infection indicators and summer mortality spikes during periods of high temperature. Influenza-specific SARI hospitalization incidence was available from 2021 onward and is shown for contextual comparison. Its peaks coincided with several prominent non-COVID SARI and mortality peaks, particularly during the winters of 2022–2025. Because influenza incidence was not used as a separate model input, this temporal agreement does not constitute a separate estimate of influenza-attributable mortality. During the pandemic phase of 2020–2022, the estimated non-COVID SARI component was lower in winter. This pattern is consistent with surveillance reports from multiple countries showing reduced circulation of non-SARS-CoV-2 respiratory viruses, including influenza and RSV, during the COVID-19 pandemic, followed by renewed winter peaks after 2022 [27]. COVID-19-associated SARI was initially concentrated in colder months, but later became less confined to winter, with activity also extending into summer 2022 and early summer 2023 [28].

The lower panel of Figure 1 shows model-estimated heat- and cold-associated deaths together with weekly and nationally averaged temperature records. Consistent with previous temperature-attribution studies, the cumulative model-estimated cold-associated component was larger than the model-estimated heat-associated component [7, 29]. This reflects the long duration of temperatures below the optimal-temperature range of approximately 15–20 *^◦^*C in Germany, rather than indicating that extreme cold necessarily produces the highest short-term risks. Model-estimated heat-associated deaths were more concentrated in short summer peaks, which is relevant for short-term public-health planning during heatwaves.

Across individual years, the model attributed 2,800–9,000 deaths per year (0.3–0.9% of annual deaths) to heat and 132,000–219,000 deaths per year (14.5–20.2%) to the combined SARI components, defined here as non-COVID SARI plus COVID-19-associated SARI. These values are conditional on the fitted model and baseline specification. In particular, the SARI component quantifies all-cause mortality variation assigned to SARI hospitalization indicators and should not, without additional cause-specific and individual-level analyses, be interpreted as a quantitative estimate of deaths caused by SARI.

### Temperature–mortality response and humidity modification

To examine the weather component underlying these seasonal patterns, Figure 2 presents the modeled increase in mortality risk associated with daily mean temperature under alternative model specifications and humidity conditions. In the model comparison, both the hybrid and weather-only models identified an optimum around 15–20 *^◦^*C, corresponding to no additional risk, and similar risk increases above 20 *^◦^*C. At lower temperatures, the weather-only model showed a steeper increase than the hybrid model, implying that models without respiratory-infection indicators may assign part of infection- related winter mortality to cold exposure.

Including dew-point temperature as a weather input improved the representation of hot and humid periods and made the weather-associated mortality component more interpretable. In the temperature–humidity exposure- response analysis, humidity had limited influence at low temperatures but amplified heat-associated mortality risk at higher temperatures. For example, at 25 *^◦^*C, the model without the dew-point humidity indicator predicted a 20.7% increase in mortality risk relative to the minimum-risk temperature. With the dew-point humidity indicator, the corresponding increases were 10.5%, 16.8%, 24.7%, 34.2%, and 45.2% at 40%, 50%, 60%, 70%, and 80% relative humidity, respectively.

### Adaptive baseline mortality

After estimating temperature- and respiratory-infection-associated mortality, Figure 3 compares registered deaths with the model-estimated adaptive mortality baseline. After subtracting these modeled components, the adaptive procedure produced a smoother baseline estimate that retained longer-term age- and sex-specific mortality patterns with less influence from short-term winter and summer peaks. Men had higher baseline mortality rates than women, and the relative male–female risk pattern was broadly stable across age groups. Overall, baseline mortality rates declined over the study period, with upward deviations in several age groups between 45 and 79 years during 2022–2023, after the main COVID-19 pandemic phase. Additional age groups are shown in Supplementary Figure 9.

**Figure 3.**
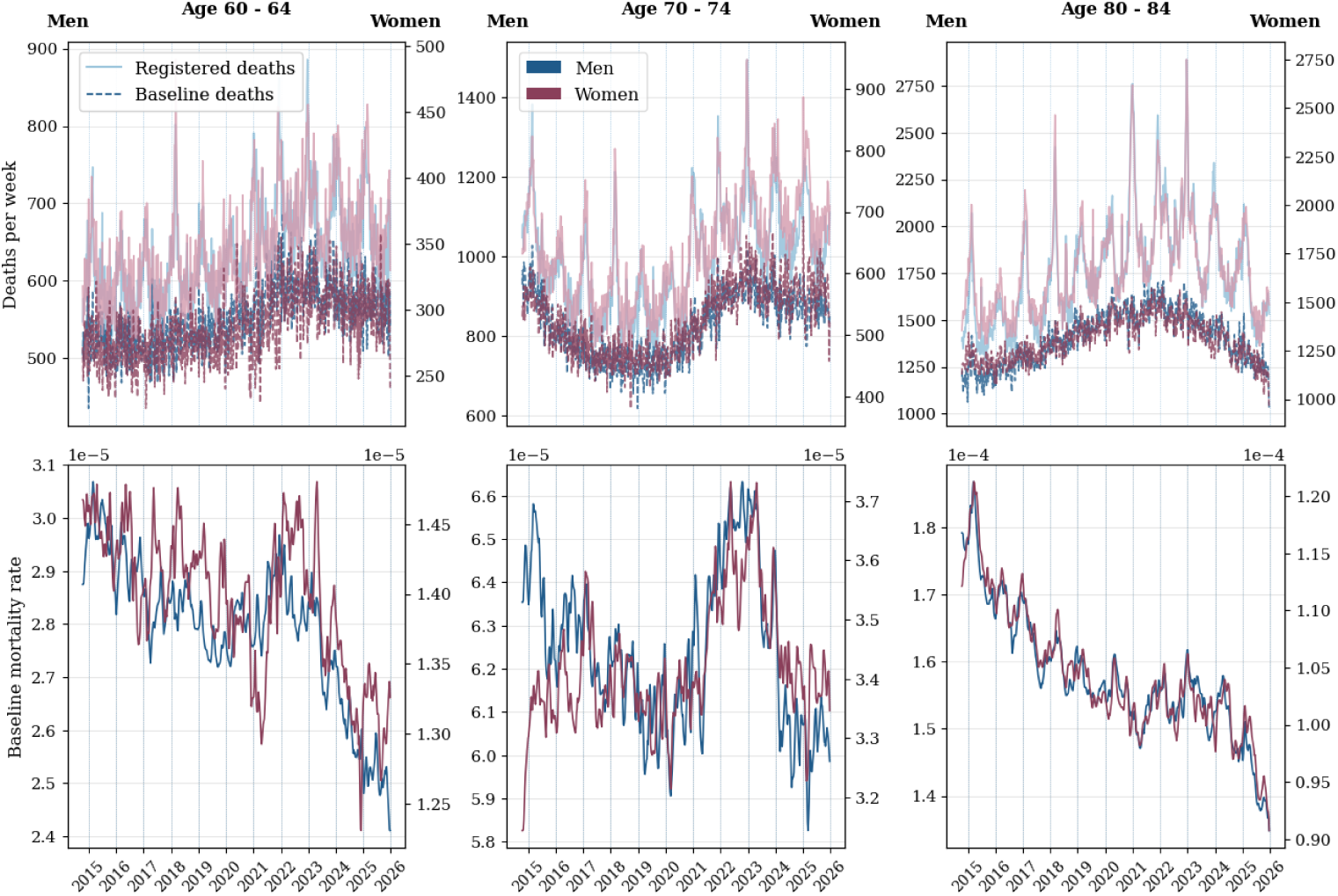
Registered deaths and adaptive baseline estimates by sex and age group, Germany, 2015–2025. Top panels show weekly registered deaths and the estimated adaptive baseline deaths. Compared with registered deaths, the adaptive baseline reduces short-term summer and winter peaks and follows the slower background mortality trend. Bottom panels show the corresponding baseline mortality rates. Women and men are shown on separate y-axis scales; in the displayed age groups, trends over time are broadly similar, while women have lower baseline mortality rates than men.

### Age- and sex-specific model-attributed mortality

To examine how model-attributed temperature- and respiratory-infection-associated deaths were distributed across demographic groups, Figure 4 summarizes average annual number of deaths and mortality rates associated with respiratory infections and extreme temperature from 2020 through 2025, stratified by sex and five-year age group. The model attributed larger absolute numbers of respiratory-infection-associated deaths than extreme-temperature-associated deaths. Below age 80, model-attributed deaths were generally higher among men; above age 85, the larger population size among women contributed to higher absolute death counts in women.

**Figure 4.**
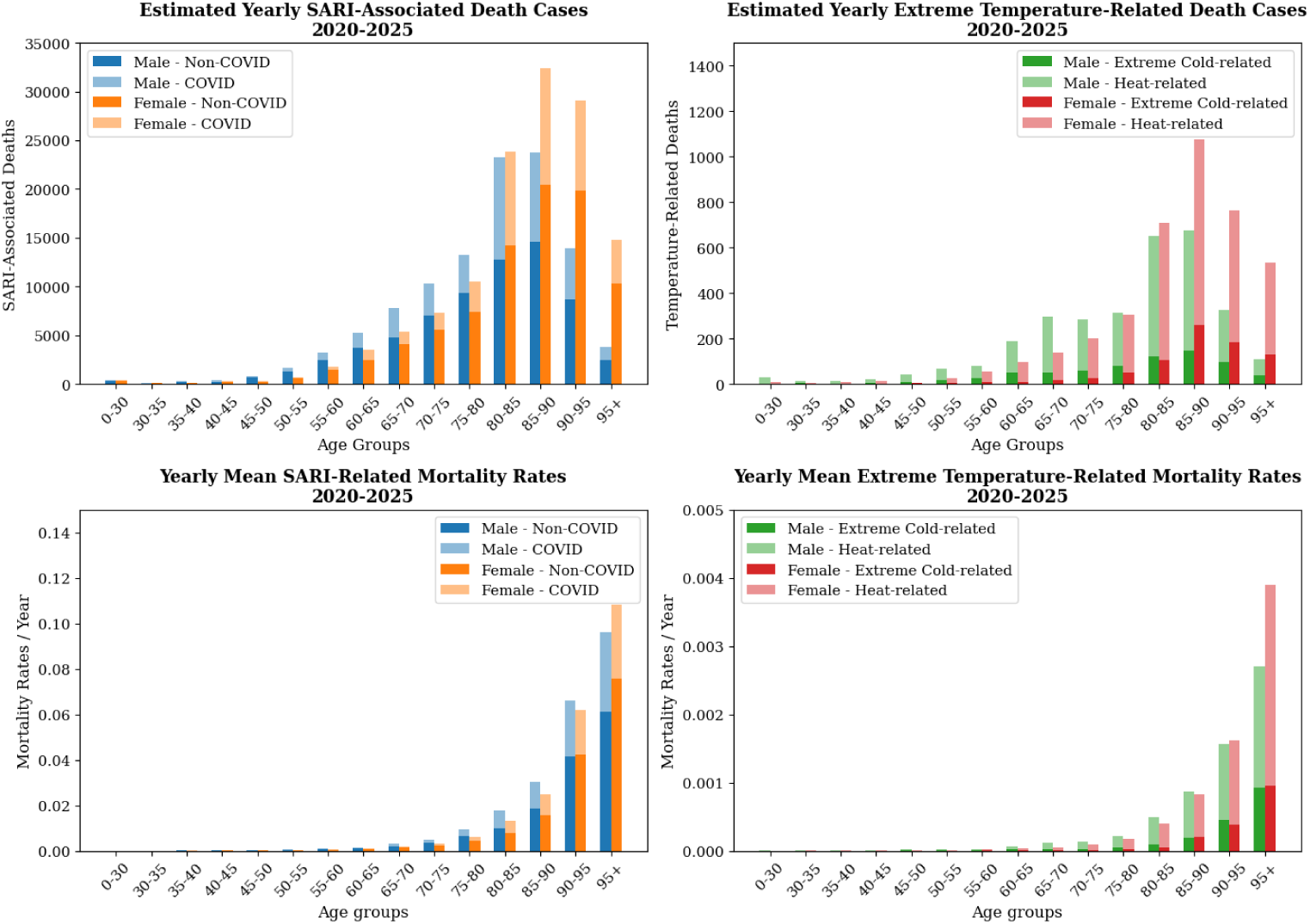
Age- and sex-specific distribution of model-attributed mortality, 2020–2025. Values represent yearly average counts and mortality rates for respiratory-infection-associated mortality (left column) and temperature-associated mortality (right column), as attributed by the fitted model. In the temperature-associated panels, heat-related deaths refer to periods above 20 *^◦^*C, and extreme-cold deaths refer to periods below 0 *^◦^*C. Independent y-axis scaling is used because the absolute magnitudes differ across components.

Population-normalized model-attributed mortality rates increased strongly with age for both respiratory-infection- and temperature-associated components. Among men, model-attributed respiratory-infection-associated mortality risks were 0.31%, 0.94%, and 3.03% at ages 65–70, 75–80, and 85– 90 years, respectively; the corresponding model-attributed female risks were 0.20%, 0.61%, and 2.48%. These patterns indicate a nonlinear age gradient and a consistent male excess in most age groups. More than 85% of model-attributed SARI- and temperature-associated deaths occurred among persons aged 70 years or older.

The age resolution of the comparison was important for interpreting sex differences in model-attributed mortality. When broad old-age categories such as 80+ years are used, the larger share of women at the oldest ages can increase the aggregate female mortality rate even if rates within narrower age bands are comparable between sexes or higher among men. The five-year age groups used here reduce this aggregation bias and show that neither respiratory-infection-associated nor temperature-associated mortality was uniformly higher among older women. The open-ended 95+ group should be interpreted cautiously because it still combines a wide age range with steeply increasing baseline mortality and may therefore retain stronger aggregation bias.

### Model performance and contribution of input components

To assess the contribution of respiratory-infection indicators, we compared model reconstructions and evaluated whether adding these indicators to weather variables improved the fit to observed all-cause mortality. As shown in Figure 5, the hybrid model integrating temperature, dew-point temperature, non-COVID SARI, and COVID-19-associated SARI achieved higher accuracy than models using only weather or respiratory-infection inputs. Across the full 2014–2025 evaluation period, after mean alignment, the ensemble median root-mean-square error (RMSE) was 80.2 deaths per day for the hybrid model, compared with 170.2 for the weather-only model and 117.1 for the variant using only non-COVID SARI and COVID-19-associated SARI inputs. Performance gains were observed across influenza-season, summer, pre-pandemic, pandemic, and post-pandemic evaluation windows (Supplementary Figure 3). The influence of other model components is shown in Supplementary Figures 4, 5.

**Figure 5.**
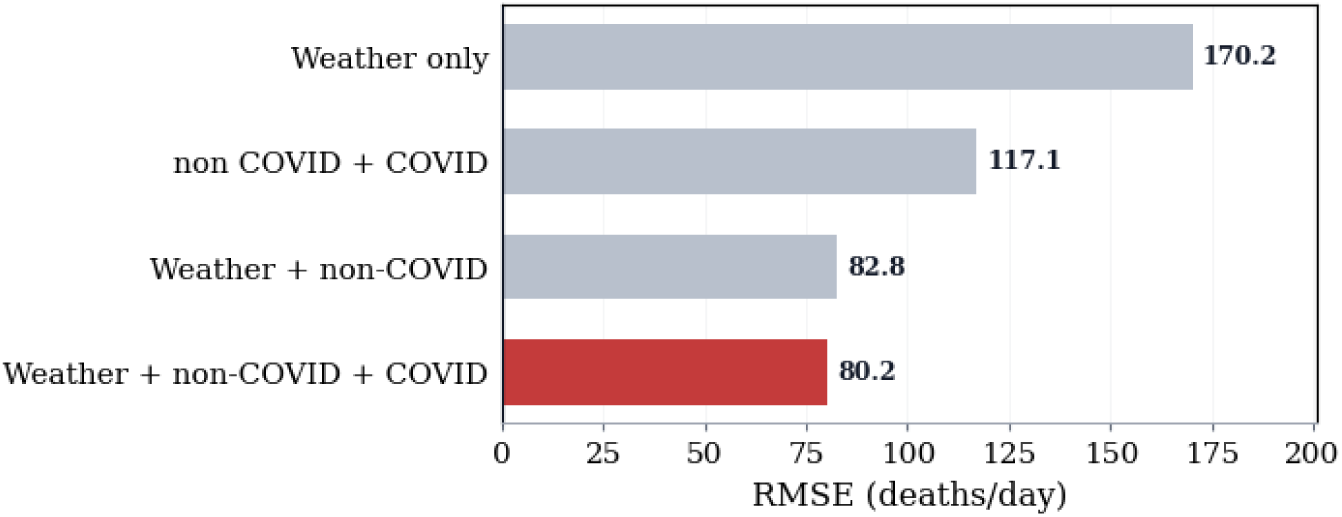
Compact comparison of model-input configurations by ensemble-median root-mean-square error (RMSE, deaths per day) over the full evaluation period. Bars are sorted from highest to lowest RMSE; lower values indicate better reconstruction of observed all-cause mortality.

As a separate robustness check for sensitivity to random initialization, 50 additional replicates of the main hybrid configuration were trained. These models achieved a mean RMSE of 80.3 deaths per day (range: 80.1–80.4), indicating stable convergence and limited sensitivity to initialization.

## Discussion

This study presents a component-based neural-network model for all-season mortality reconstruction and attribution that jointly uses weather, respiratory-infection surveillance, demographic structure, and an adaptive baseline. Combining environmental and respiratory-infection indicators allowed a consistent reconstruction of all-cause mortality dynamics in Germany, where short-term seasonal components can be examined separately from slower background mortality changes. Accordingly, the primary evaluated result is the improved reconstruction of observed all-cause mortality; the component-specific mortality estimates are interpreted as structured, model-based decompositions conditional on the fitted model rather than as a unique, directly observed partition of mortality.

This performance gain is important because it indicates that environmental and infectious-disease inputs explain complementary parts of the all-cause mortality dynamic. In an all-season setting, winter respiratory-infection peaks and summer heat events occur within the same time series, so models that omit either input class risk assigning shared seasonal variation to the remaining predictors.

Introducing respiratory-infection indicators is central to a nuanced interpretation of winter mortality attribution. Compared with a temperature-only model, the hybrid model captured part of the winter mortality signal through respiratory-infection indicators rather than assigning it to cold exposure. This interpretation is consistent with the low non-COVID respiratory infection signals during 2020–2022, when non-pharmaceutical interventions suppressed circulation of several respiratory viruses [30, 31]. The finding should be interpreted as evidence for shared winter drivers rather than as a claim that cold exposure is unimportant. Temperature remained an important component, but the comparison with the temperature-only model suggests that direct cold effects and infection-mediated winter mortality may be difficult to separate without infection indicators.

The component-specific outputs are supported by plausibility and external- consistency checks rather than by direct validation against cause-specific death records. For temperature-associated mortality, the fitted response showed a minimum around 15–20 *^◦^*C and stronger risk under hot, humid conditions, consistent with established temperature–mortality evidence and physiological expectations about humid heat stress [7, 32]. The model- attributed heat-related burden showed broadly similar year-to-year variation to published German heat-mortality estimates, while differences in study period, baseline definition, exposure metric, spatial resolution, and attribution threshold affect absolute numbers (Supplementary Figure 6). For respiratory-infection-associated mortality, the fitted component was compared with broader SARI-based and IQM any-mention estimates as well as official primary-cause death counts (Supplementary Figures 7 and 8) [17, 33, 34, 35]. These comparisons provide plausibility checks on temporal patterns and relative magnitudes, but they do not validate the component totals as causal estimates or make them directly interchangeable with surveillance- or death-certificate-based categories.

The interpretation of these components also depends on how the background mortality level is defined. The adaptive baseline is therefore an additional methodological contribution. Its main value is not that it provides a definitive exposure-free baseline, but that it offers a transparent residual-based approach to separate short-term modeled deviations from slower background mortality changes. Conventional baseline estimation can be difficult when reference periods themselves contain exposure-related mortality, as in severe winters, heatwave periods, or summers with unusual respiratory activity [15]. Because winter respiratory-infection peaks are modeled separately, the adaptive baseline is less likely to treat them as part of background mortality. The residual-based adaptive baseline improved reconstruction accuracy and may provide a practical alternative in periods where conventional anchor periods were contaminated, especially during the COVID era. The resulting age- and sex-specific baseline trajectories may also be relevant for demographic or epidemiologic research because they can flag unusual background-mortality shifts and support hypothesis generation, although they do not by themselves explain the causes of those shifts.

Beyond the seasonal attribution, the fitted components also showed strong demographic gradients and clarified an important interpretation issue for age- and sex-specific mortality attribution. Previous studies have often reported higher heat-related mortality among older women[12], but similar aggregation problems can affect both temperature- and respiratory-infection- associated mortality. In particular, grouping all persons aged 80 years and older can produce a Simpson’s-paradox-like pattern: because women make up a larger share of the oldest-old population, the aggregate female rate can be higher even when rates within narrower age bands are similar between sexes or higher among men. By using five-year age groups and population- normalized rates, our analysis reduces this aggregation bias and suggests that apparent female excess in either temperature- or respiratory-infection- associated mortality should not be interpreted without attention to the underlying age structure. The open-ended 95+ category remains an exception to this fine-stratification strategy and should be interpreted cautiously, because age composition within this group can still differ by sex and may retain the same aggregation bias.

Several limitations affect interpretation of the attribution framework and component estimates. Most importantly, the annual totals assigned by the model to the SARI components are not quantitative estimates of deaths caused by SARI. They represent the amount of all-cause mortality variation assigned to SARI hospitalization indicators within the fitted decomposition. Because the model was trained on all-cause mortality without individual infection status, causes of death, contributory conditions, or underlying diseases, it cannot determine whether an infection caused, precipitated, or coincided with a death, and the fitted components may also capture unmeasured factors that vary with SARI activity. The model also assumes time-invariant relationships between SARI hospitalization incidence and mortality for non-COVID SARI and COVID-19-associated SARI. This is a reasonable first approximation for non-COVID respiratory infections but is likely to over- simplify COVID-19 dynamics, where severity varied by variant, immunity, treatment, and vaccination status [36]. In addition, the non-COVID SARI component combines influenza with other non-COVID respiratory infections and cannot be decomposed by pathogen in the present analysis. Future work could extend this decomposition to influenza, RSV, and other respiratory pathogens when longer, consistently defined pathogen-specific surveillance series become available. Furthermore, SARI hospitalization data are available only at the national level, so regional heterogeneity in infection pressure cannot yet be modeled. Finer-scale respiratory-infection surveillance would improve spatial attribution and reduce reliance on national incidence assumptions. Finally, the study does not report conventional confidence intervals for all component-attributable estimates. This does not imply that the estimates are treated as certain. Rather, conventional likelihood-based intervals conditional on a single fixed model could be misleading, because they would capture only part of the uncertainty while excluding major sources of structural uncertainty in attribution. These include baseline specification, surveillance measurement error, lag and smoothing choices, and residual temporal dependence. Where possible, we therefore report ensemble-based ranges and sensitivity analyses to assess the robustness of the estimated components across plausible model specifications.

These data limitations also shape how the framework could be generalized. Although this application uses structured SARI hospitalization data, the broader modeling approach could be explored in settings where infection indicators are incomplete. Whether grouped cause-of-death information or other respiratory-infection proxies can provide useful partial infection signals should be evaluated in future applications before the framework is generalized beyond settings with comprehensive SARI hospitalization surveillance.

If supported by suitable surveillance data, this type of integrated attribution framework may provide evidence relevant to vaccination strategy, respiratory-virus surveillance, heat-health planning, and hospital-capacity management. In Germany, influenza-vaccination coverage remains below the EU target of 75% [37], with coverage among adults *≥*60 years around 45% in 2023 and below 25% among those with chronic disease [38]. The present model does not directly evaluate intervention effects, but by estimating respiratory-infection, temperature, and baseline components within one model it may support prioritization, monitoring, and hypothesis generation for prevention strategies.

Looking ahead, longer and more severe heatwaves [39], potential shifts in respiratory-infection seasonality [40], and demographic change increase the need for monitoring systems that can integrate environmental and infectious drivers of mortality [41, 40]. The framework presented here provides methodic basis for such integration and could support future mortality surveillance, early-warning systems, and comparative analyses across populations where suitable surveillance data are available.

## Contributors

J.W., M.a.d.H., and C.I. conceived the study. J.W. collected, processed, and validated the data. J.W. and C.I. designed the model. J.W., M.a.d.H., and C.I. conducted the investigation and interpreted the results. J.W. and C.I. wrote the original draft. All authors reviewed and edited the manuscript. C.I. supervised the study.

## Data sharing statement

The mortality and population data used in this study are publicly available from the Federal Statistical Office of Germany and the Regional Database Germany [21, 22]. HOSTRADA weather data are available from the German Weather Service [23], and the SARI hospitalization-incidence data are available through Zenodo [25]. Processed data and fitted model parameters are available at https://github.com/ClimSocAna/sari-weather-mortality/.

## Code availability

All code used for data processing, model fitting, evaluation and figure generation is publicly available at https://github.com/ClimSocAna/sari-weather-mortality/.

## Data Availability

All data produced are available online at

https://github.com/ClimSocAna/sari-weather-mortality/

## Acknowledgements

The Federal Ministry of Research, Technology and Space (BMFTR) supported this study through funding for the CLIMADEMIC project (funding code 01LN2210A) within the Research for Sustainability (FONA) strategy. We thank Walter Haas, Udo Buchholz and Kristin Tolksdorf from Unit 36, Respiratory Infections, Department of Infectious Disease Epidemiology, Robert Koch Institute, for providing the SARI data and for constructive discussions and feedback that improved the manuscript.

