## Supplementary for "Integrating respiratory infection surveillance and temperature data improves all-season mortality reconstruction"

### Supplementary figures

### Supplementary Figure 1

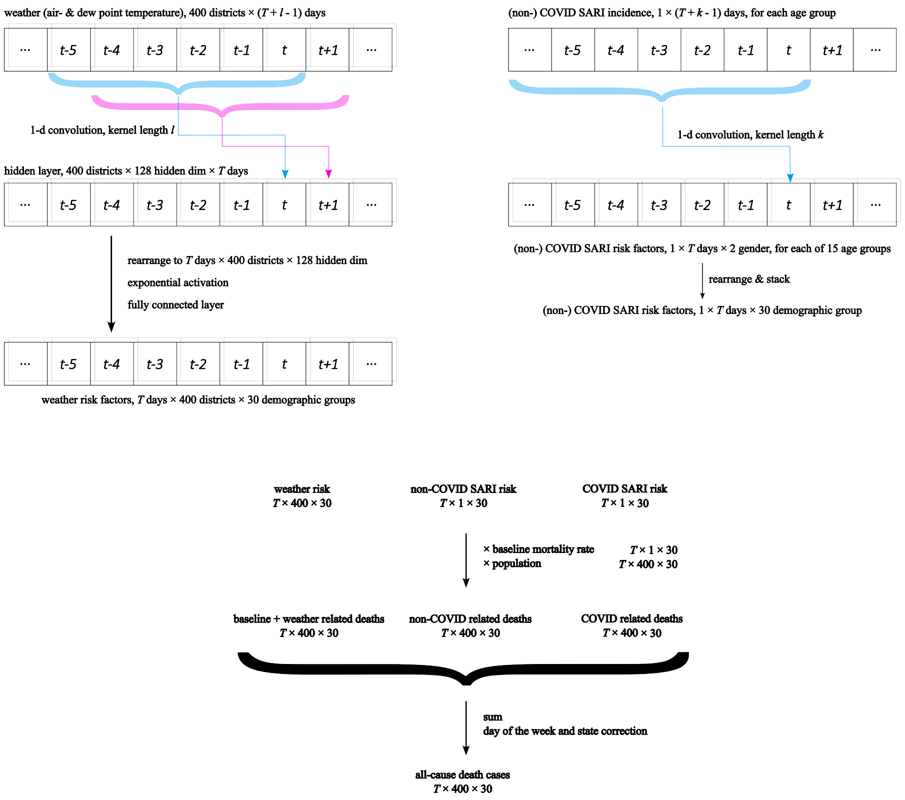


Supplementary Figure 1: Network architecture of the component-based neural-network model used to estimate daily all-cause mortality. The model combines weather indicators, non-COVID SARI, and COVID-19-associated SARI inputs with an adaptive baseline mortality component.

### Supplementary Figure 2

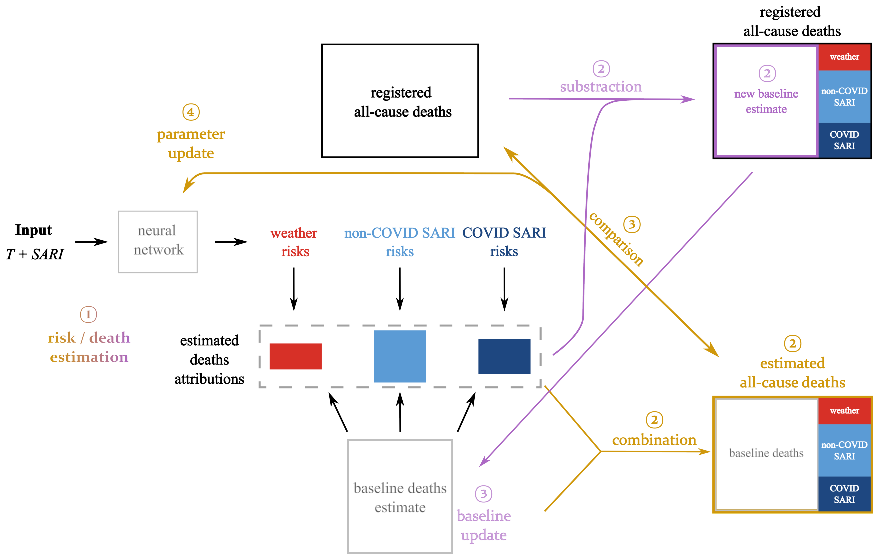


Supplementary Figure 2: Training workflow for the mortality-attribution model. Yellow arrows indicate model-training and parameter-update steps, while pink arrows indicate adaptive baseline updates.

### Supplementary Figure 3

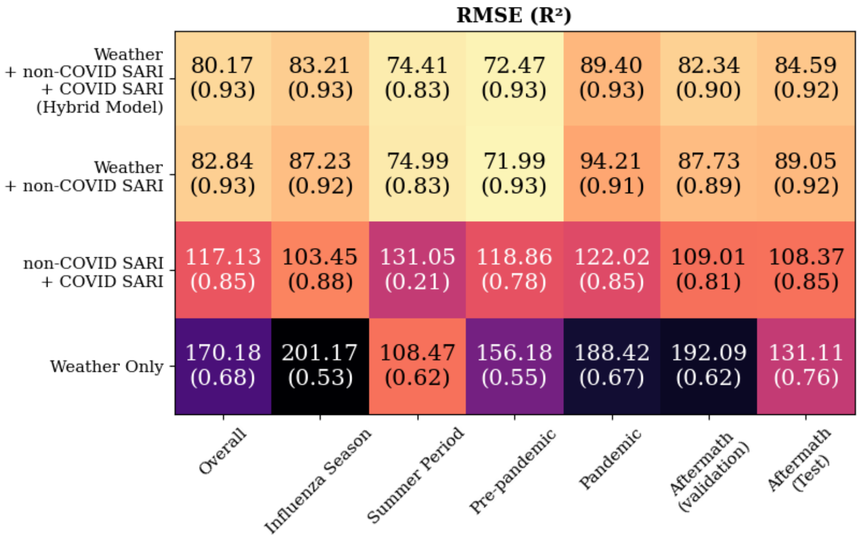


Supplementary Figure 3: Model-performance comparison across input configurations, using the mean-aligned Poisson training objective. Heatmaps show RMSE and $R^2$ for models using weather + non-COVID SARI + COVID-19-associated SARI, weather + non-COVID SARI, non-COVID SARI + COVID-19-associated SARI only, and weather only. Performance is shown for evaluation windows covering the full 2014–2025 evaluation period, influenza season (week 40 to week 20), summer period (week 15 to week 38), pre-pandemic period (through 2019), pandemic period (2020–2022), and post-pandemic period (2023–2025, including the 2023–2024 validation portion and the 2025 test period). Weather inputs include air temperature and dew point; the baseline adaptation method is median-EWMA.

### Supplementary Figure 4

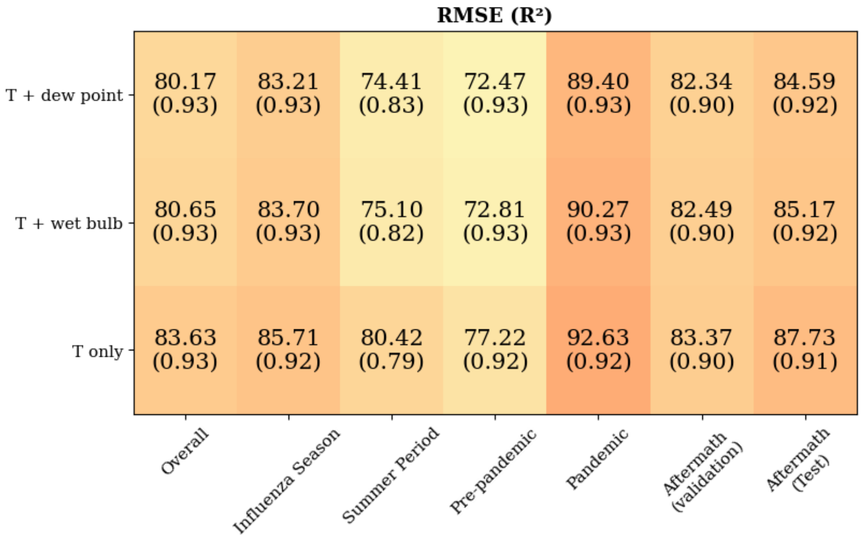


Supplementary Figure 4: Sensitivity of model performance to weather-input specification, using the mean-aligned Poisson training objective. Heatmaps show RMSE and $R^2$ for models using air temperature + dew point, air temperature + wet-bulb temperature, and air temperature alone across the same evaluation windows as in Supplementary Figure 3. All configurations include non-COVID SARI and COVID-19-associated SARI inputs and use the median-EWMA baseline adaptation method.

### Supplementary Figure 5

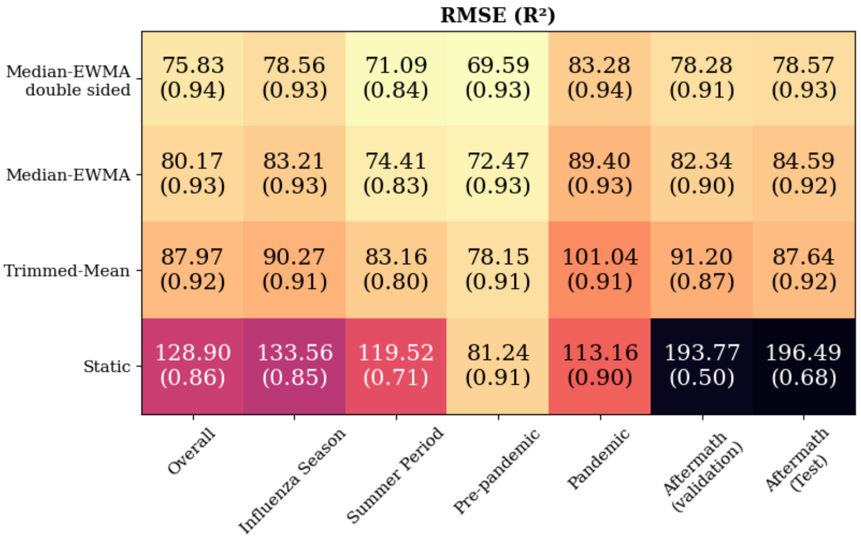


Supplementary Figure 5: Sensitivity of model performance to baseline-adaptation strategy, using the mean-aligned Poisson training objective. Heatmaps show RMSE and $R^2$ for double-sided median-EWMA, single-sided median-EWMA, trimmed-mean, and static baseline strategies across the same evaluation windows as in Supplementary Figure 3. Weather inputs are air temperature and dew point; respiratory inputs are non-COVID SARI and COVID-19-associated SARI.

### Supplementary Figure 6

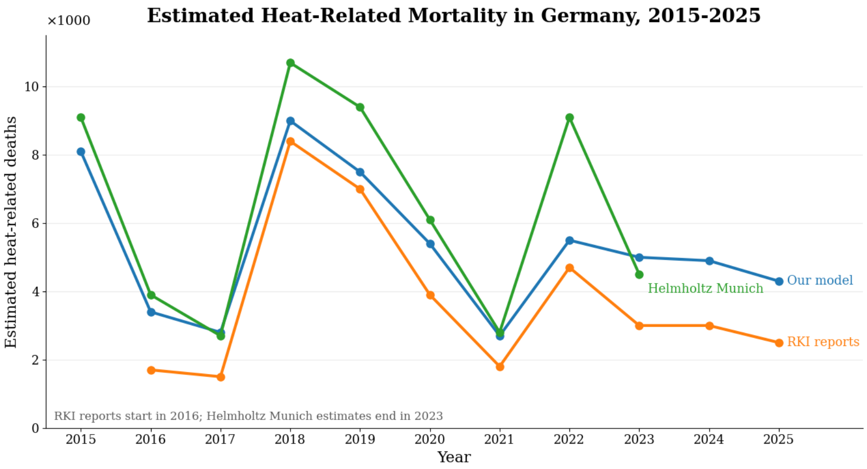


Supplementary Figure 6: External comparison of annual heat-related mortality estimates in Germany, 2015–2025. The hybrid-model estimates are compared with published RKI heat-mortality estimates[1] and Helmholtz Munich estimates[4] where available. Differences between series should be interpreted in light of differences in study period, exposure definition, baseline construction, spatial resolution, and attribution threshold.

### Supplementary Figure 7

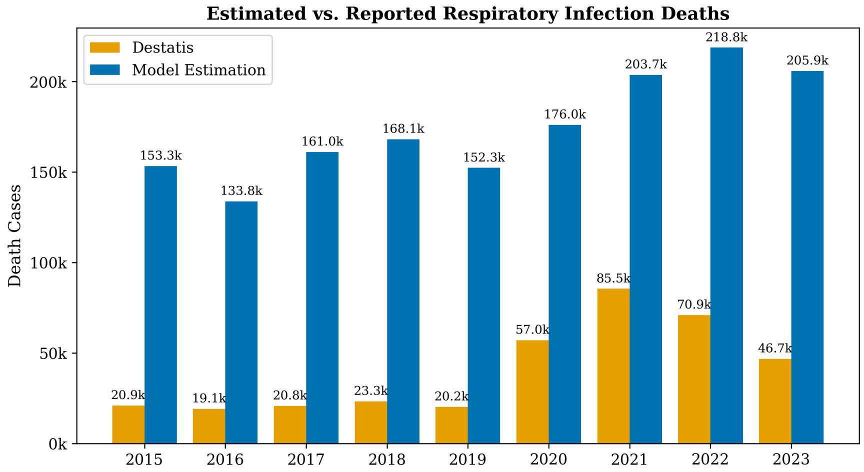


Supplementary Figure 7: External comparison of respiratory-infection mortality estimates in Germany, 2015–2023. The figure compares official primary-cause counts from Destatis[3] with hybrid-model estimates. Destatis categories include TDU-081 (influenza), TDU-082 (pneumonia), and TDU-18/19 (COVID-19 with or without virus detection). This comparison is intended as a contextual plausibility check and should be interpreted in light of differing case definitions and attribution rules.

### Supplementary Figure 8

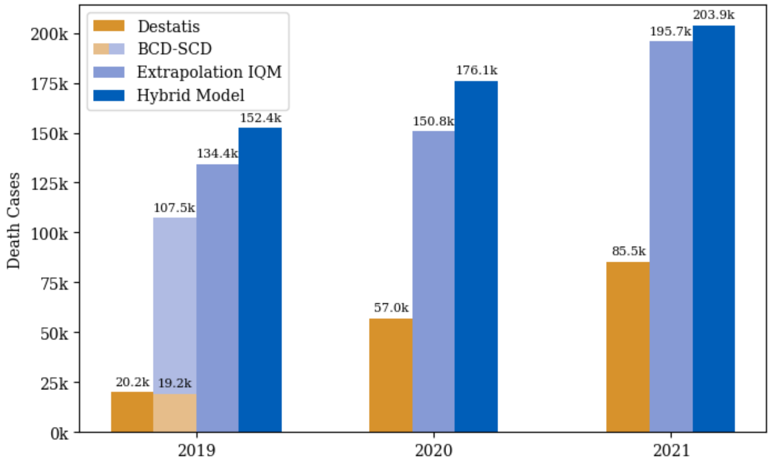


Supplementary Figure 8: Focused comparison of respiratory-infection mortality estimates for 2019–2021. Bars compare official primary-cause counts from Destatis,[3] BCD–SCD estimates,[2] IQM extrapolations,[5,6] and the hybrid-model estimates. The comparison illustrates how model-attributed respiratory-infection mortality relates to narrower cause-of-death counts and broader hospitalization-based estimates during the pre-pandemic and early pandemic years.

### Supplementary Figure 9

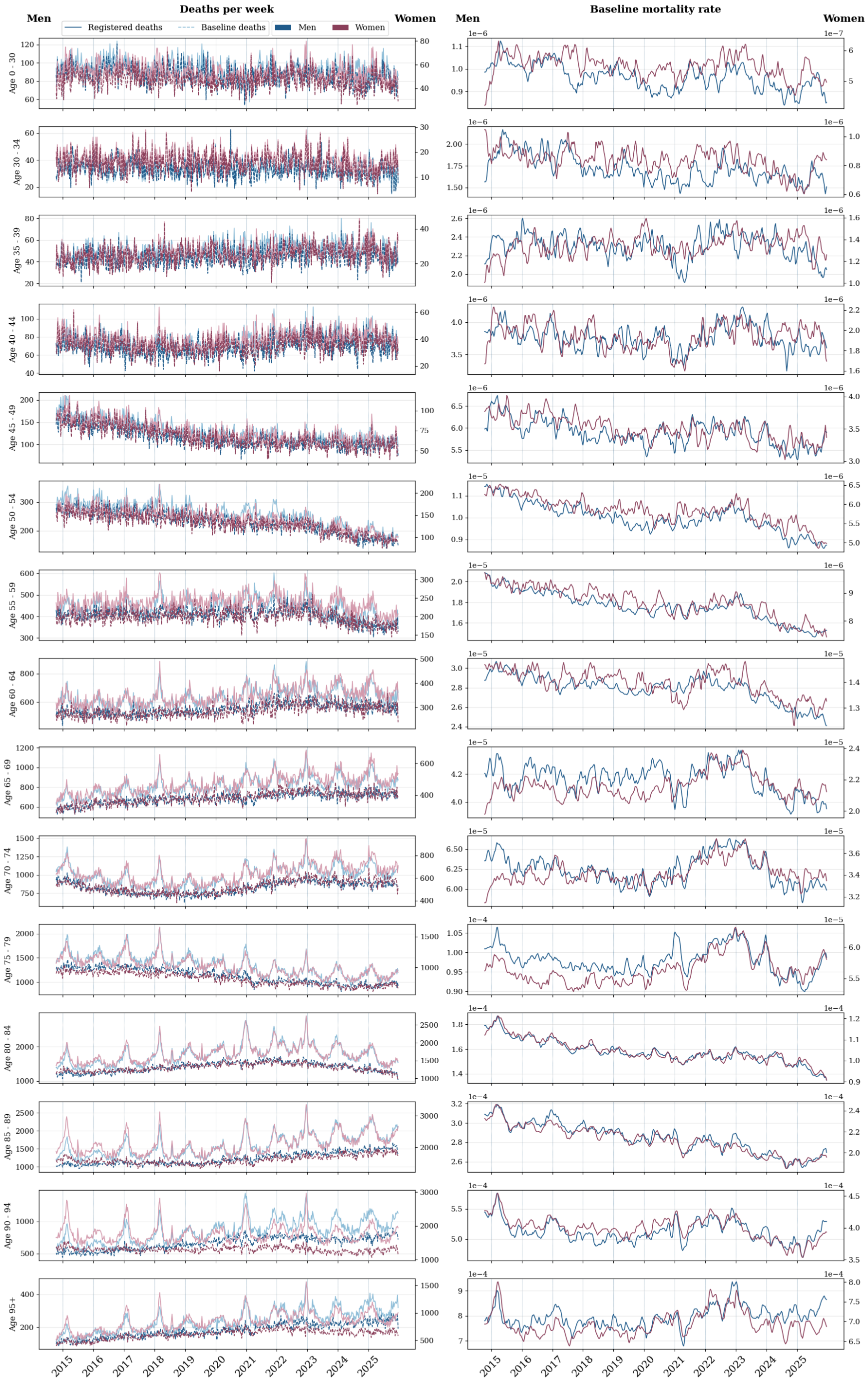


Supplementary Figure 9: Weekly registered deaths and estimated baseline deaths by age group and sex, Germany, 2015–2025. Left panels show registered and estimated baseline deaths; right panels show the corresponding baseline mortality rates.

### Supplementary References

[1] Matthias an der Heiden, Benedikt Zacher, RKI-Geschäftsstelle für Klimawandel & Gesundheit, Michaela Diercke, and Viviane Bremer. Wochenbericht zur hitzebedingten mortalität kw 38/2025 vom 02.10.2025, 2025.

[2] Silke Buda, Kristin Tolksdorf, Ekkehard Schuler, Ralf Kuhlen, and Walter Haas. Establishing an icd-10 code based si-surveillance in germany–description of the system and first results from five recent influenza seasons. *BMC public health*, 17(1):612, 2017.

[3] Federal Statistical Office of Germany (Destatis). Deaths: Germany, years, causes of death, sex, 2025.

[4] Veronika Huber, Susanne Breitner-Busch, Hanna Feldbusch, Katja Frieler, Cheng He, Franziska Matthies-Wiesler, Matthias Mengel, Siqi Zhang, Annette Peters, and Alexandra Schneider. Improvements in life expectancy mask rising trends in heat-related excess mortality attributable to climate change. *Nature Communications*, 2025.

[5] Initiative Qualitätsmedizin. Covid-19 pandemie, 2021.

[6] Initiative Qualitätsmedizin. Effekte der sars-cov-2 pandemie auf die stationäre versorgung 2020., 2021.
